# Evaluating menstrual-friendly public bathrooms in Atlanta, Georgia: An audit of availability, accessibility, and resources

**DOI:** 10.64898/2026.11.11.26367812

**Authors:** April M. Ballard, Emily Wallace, Brandie Banner Shackelford, Marcus Cain, Adriana Williams, Thomas J. “Chip” Daymude, Shannon Self-Brown, Christine Stauber

**Affiliations:** Department of Population Health Sciences, School of Public Health, Georgia State University; Center on Health and Homelessness, Georgia State University; Department of Health Policy & Behavioral Sciences, School of Public Health, Georgia State University; Gangarosa Department of Environmental Health, Rollins School of Public Health, Emory University

## Abstract

Public toilets are essential urban infrastructure that enable people who menstruate to participate in public life with dignity and comfort. However, little is known about whether publicly accessible toilets adequately support menstrual health and hygiene, particularly in high-income urban settings where privately operated facilities often function as de facto public infrastructure. This study assessed the menstrual friendliness of publicly accessible toilets in Atlanta, Georgia. We also examined differences by funding source (i.e., government versus private) and neighborhood type. We conducted field-based audits of women’s, gender-neutral, and family toilets across 15 areas. Using an adapted Menstrual Friendly Public Toilet framework, we evaluated six dimensions: cleanliness, privacy and safety, structural features, general resources, accessibility, and menstrual health and hygiene resources. We calculated domain-specific and overall scores and compared government- and privately funded facilities, with additional descriptive comparisons across business, residential, tourist, and transit neighborhoods. We audited 125 toilet facilities across 112 locations. The overall mean Menstrual Friendly Public Toilet Index was 0.797. Cleanliness (0.974), privacy and safety (0.954), structure (0.930), and general resources (0.899) scored relatively high, whereas accessibility (0.622) and menstrual health and hygiene resources (0.490) scored lowest. Only 4.0% of facilities provided menstrual products or a functional dispenser. Overall scores did not differ significantly by funding source, although government-funded facilities had higher structural feature scores than privately funded facilities (0.952 versus 0.913; p=0.027). Menstrual health and hygiene resource scores varied across neighborhood types, ranging from 0.200 in the residential area to 0.775 in the tourist area. These findings demonstrate that the availability of clean and functional bathrooms does not necessarily ensure people can fully meet and manage their menstrual needs. Addressing gaps in accessibility and menstrual-specific resources will require coordinated attention to both government- and privately managed facilities, as well as consideration of neighborhood context to support menstrual health and equitable participation in urban public spaces.

## Introduction

Access to safely managed and adequate household sanitation is a fundamental public health need, yet 3.4 billion individuals globally still lack access (1, 2). As a result, many rely on public or community-based sanitation facilities as critical access points. Public facilities can promote health, dignity, and equitable participation in public spaces by enabling individuals to meet their biological needs without restricting movement through transit systems, workplaces, and other community environments (3–7). However, public sanitation systems remain unevenly distributed and frequently under-resourced, reflecting competing priorities related to costs, safety, and governance of public spaces despite their importance (7–13). These dynamics shape sanitation access, as well as who can participate in everyday public life without compromising their health, comfort, and dignity.

Menstrual health and hygiene (MHH) is a critical yet frequently neglected dimension of sanitation, requiring public bathrooms that support safe, dignified, and inclusive menstrual management (14–18). Approximately one in three women and girls lack access to adequate sanitation facilities, limiting their ability to manage menstruation safely and hygienically (2). Insufficient sanitation access during menstruation can result in psychosocial distress, anxiety, and increased vulnerability to gender-based violence, while also limiting participation in education and employment opportunities (2, 18–20). Inadequate public sanitation infrastructure can further exacerbate stigma surrounding menstruation, reinforcing experiences of exclusion and discomfort in public spaces (17, 21, 22). These challenges disproportionately affect those who menstruate and reflect broader gendered inequities in access to public space and urban infrastructure (13, 15, 18, 23). Still, MHH needs are rarely prioritized in the design, provision, or evaluation of public sanitation systems.

The concept of Menstrual Friendly Public Toilets (MFPT) was introduced through a global multi-city audit to highlight the need for public sanitation systems that better support those who menstruate (14, 24). The MFPT framework evaluates facilities across key domains, including accessibility, privacy and safety, structure, general resource availability, cleanliness, and MHH-specific supports. Findings from the initial MFPT study across diverse cities demonstrated that there is a significant gap in the provision of MFPTs in urban areas (14). While this study significantly characterized MFPTs across six cities, important gaps remain in understanding how these features vary across other contexts.

Over 99% of the US population uses at least basic household sanitation facilities (i.e., use of improved facilities [e.g., flush/pour flush to piped sewer systems, septic tanks or pit latrines] that are not shared with other households), with most (97%) having the safest type of sanitation (i.e., safely managed sanitation defined as the use of improved facilities that are not shared with other households and where excreta are safely disposed of in situ or transported and treated offsite) (1, 25, 26). Conversely, access to public sanitation infrastructure (i.e., public bathrooms) remains limited, fragmented, and inconsistently governed (7–9, 24, 27, 28). Public facilities are often concentrated in specific locations such as parks and tourist destinations, while many urban areas increasingly rely on privately funded facilities (e.g., restaurants, retail businesses) to meet public sanitation needs (8, 9, 28), especially as government investment in sanitation infrastructure has declined. Access to these private facilities, however, is often conditional and shaped by social and economic norms, creating barriers for individuals without purchasing power or perceived legitimacy as customers (8, 9). These structural dynamics may contribute to uneven access to sanitation and further limit the ability to manage menstruation in public spaces. In recognition of this reality, we employ an inclusive operational definition of *public* bathrooms that encompasses both government-funded and privately funded facilities in this study. Additionally, because the MFPT framework was originally developed to assess publicly accessible sanitation facilities across diverse contexts, the prominence of privately funded facilities in the US presents an opportunity to examine how the framework performs in settings where access to public sanitation is frequently mediated through private ownership and consumer-based norms.

Atlanta, Georgia provides a relevant case study for examining these dynamics in a high-income urban context. The city reflects broader US patterns, including a mix of government-funded and privately funded facilities that serve public users, alongside ongoing challenges related to access, maintenance, and equitable distribution (29). Building on evidence from other urban areas in the US, including New York City (14, 24), San Diego (30, 31), and Saint Louis (32), Atlanta offers an opportunity to examine how public sanitation systems function in practice and how they support (or fail to support) MHH and inclusion in everyday environments. Specifically, this study evaluates the menstrual friendliness of publicly accessible bathrooms in Atlanta, Georgia. Using the MFPT framework (14), we (1) quantify overall menstrual friendliness and key MFPT sub-domains, (2) examine differences by funding type (government versus privately funded facilities), and (3) assess variation across neighborhood types. Through this study, we also explore the applicability of the MFPT framework within a US public-private sanitation landscape, expand the global evidence base, and bridge this global measurement approach with local realities. The findings presented here can be used to inform policy development, urban design, and public health programming aimed at promoting inclusive sanitation and advancing gender equity in public spaces.

## Methods

We conducted field-based audits of government-funded and privately funded bathroom facilities between February and April 2025 to evaluate the menstrual friendliness of public bathroom facilities in Atlanta, Georgia. *Government-funded facilities* included those partially or fully supported by federal, state, or local public entities and theoretically open to the public (e.g., government buildings, police stations, universities). *Privately funded facilities* included locations commonly used as de facto public bathrooms in the US (e.g., gas stations, fast-food chain restaurants, chain coffee shops, grocery stores). Private funded facilities were selected based on national presence to enhance generalizability.

### MFPT audit tool

We adapted the MFPT Tool, originally developed to assess “menstrual friendliness” of public bathrooms across multiple countries, including the US, Brazil, Spain, Japan, Uganda, and the Philippines (14). The tool standardizes data collection, including basic information (e.g., hours of operation), accessibility (e.g., permission required to use toilet), structure (e.g., functional locks), sink and toilet functionality (e.g., quantity), cleanliness (e.g., odor), and availability of menstrual management supplies and hygiene items (e.g., menstrual materials, disposal bins, soap, toilet paper). The revised tool was programmed in Qualtrics (Provo, UT, USA) and consisted of two forms. The first form (≤32 items) captured location-level characteristics, including building type, accessibility, geolocation, and hours of operation. The second form (≤86 items) captured detailed information for each bathroom type (i.e., women’s, gender-neutral, and family). Skip logic reduced respondent burden, resulting in 19 to 118 items per audit. The average completion time was 17.4 minutes (median = 8.6 minutes). Although men’s bathrooms were also audited for the broader study, those results are reported elsewhere (29).

### Adaptions to MFPT Indicators

To improve contextual relevance and data quality, we made several modifications to the original MFPT Tool. First, bathrooms with “no door by design” were scored as 0.5 rather than 0 to reflect partial privacy as opposed to complete absence. Second, for stall-level indicators, denominators were restricted to unoccupied and accessible stalls to avoid penalizing facilities where some stalls could not be assessed. This approach was applied to indicators including functional locks, functional doors, hooks or shelves, toilet paper, and disposal bins. Third, functional hand sanitizer was accepted as an alternative to soap when assessing hand hygiene resources. Fourth, we removed ambiguous language (e.g., “other materials”) from cleanliness indicators to improve clarity. Fifth, we expanded response options for menstrual-specific disposal bins to capture whether bins were present, lined, and lidded. Sixth, mirror criteria were refined to reflect adequate clarity and length for clothing checks. The full audit tool is provided in the Supplementary Material.

### Data collection

Audits were conducted across 15 distinct areas (Figure 1, Panel A) during weekday, non-holiday business hours (8AM-5PM). Audit areas were selected based on high pedestrian traffic and the presence of key features (e.g., tourist destinations, transit centers) or populations (e.g., areas frequented by people experiencing homelessness). Following the MFPT audit protocol (14), we defined each area using a 500-meter radius buffer around a central landmark (e.g., transit stations, large parks, downtown areas) using ArcGIS Online (version 2024.3), adjusting buffers to prevent overlap.

**Figure 1.**
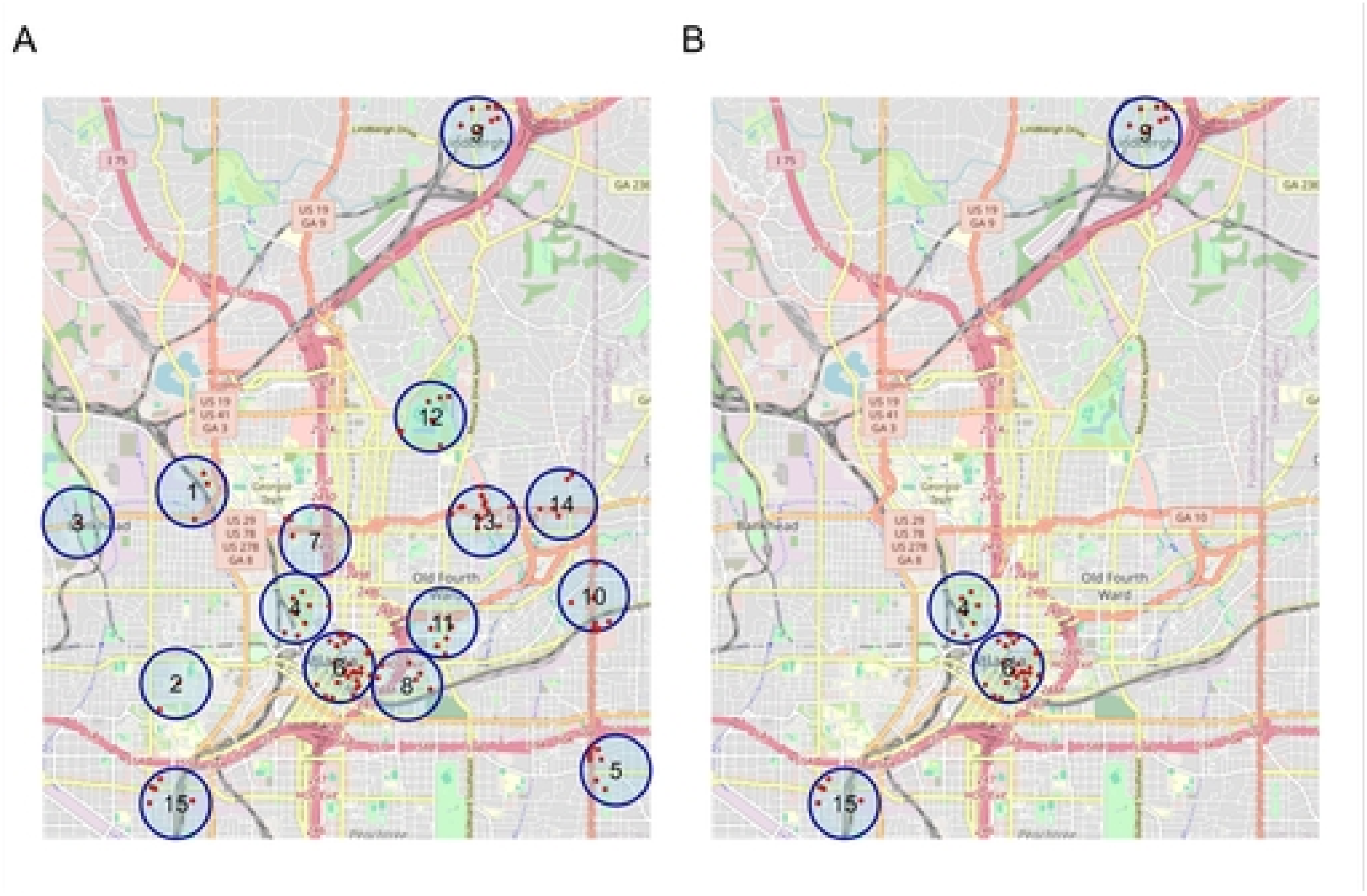
Audited locations and 500-meter buffers in a 2025 audit of public toilets in Atlanta, Georgia, USA with red pins marking audited locations. Panel A includes the entire sample. Panel B includes facilities in the following neighborhood types: 1) business area, 2) transit hub, 3) tourist area, and 4) working-class residential neighborhoods (residential). Basemap tiles from OpenStreetMap contributors, accessed via the basemaps R package. *Panel B: #4 – Tourist area, #6 – Business area, #9 – Transit hub, #15 – Working-class residential neighborhood

Research team members (MC, AW, TD, ME, BBS) were trained to identify eligible locations by systematically walking all pedestrian-accessible streets within each buffer in teams of two or three. Upon identifying an eligible location, team members jointly completed the first form and then independently completed the second form for each available bathroom type (i.e., women’s, men’s, gender-neutral, and family/caregiver). When multiple bathrooms of the same type were available, the facility closest to the main building entrance was audited. If a bathroom was occupied, team members waited 10 minutes before proceeding.

### Statistical Analysis

First, we calculated descriptive statistics to characterize the audited locations and bathroom facilities. Second, to assess menstrual friendliness, we calculated sub-domain indices and an overall MFPT Index score for each facility. The MFPT Index, as conceptualized in the muti-city audit (14), includes six sub-domains comprising 28 variables: 1) accessibility, 2) structural privacy/safety, 3) structure, 4) availability of general resources, 5) cleanliness, and 6) availability of MHH resources. All variables were dichotomized to reflect desirable MFPT characteristics (coded as 1 = present). Count variables were converted to threshold indicators. For example, the number of stalls with functional locks was recoded to indicate whether ≥75% of assessed stalls had functional locks.

Sub-domain indices ranged from 0 to 1 and represented the proportion of criteria met. For instance, the structural privacy/safety sub-domain included four indicators: (1) functional entrance door or no door by design, (2) ≥75% unoccupied stalls with functional doors, (3) ≥75% of unoccupied stalls with functional locks, and (4) functional electric light. A facility meeting 3 of 4 criteria would have a domain score of 0.75 [(1+1+1+0)/4]. The overall MFPT Index was calculated as the mean of the six sub-domain scores, where values closer to 1 indicate that facilities had a relatively higher level of “menstrual friendliness.”

Third, we stratified analyses to compare government-funded and privately funded facilities and their menstrual friendliness. We performed t-tests to compare continuous MFPT sub-domain and overall Index scores and χ^2^ tests to compare categorical variables. Statistical significance was defined as α = 0.05.

Fourth, we restricted the dataset to only include facilities in the following neighborhood types: 1) business area, 2) transit hub, 3) tourist area, and 4) working-class residential neighborhoods (residential). These neighborhood types were defined based on the multi-city MFPT criteria (14) and were each restricted to one of the 15 audit areas for a total of four areas (Figure 1, Panel B). Descriptive statistics were calculated to show the overall characteristics of facilities across all four neighborhoods and stratified by neighborhood type to compare differences.

Finally, we conducted sensitivity analyses using the entire dataset to assess the potential influence of bathroom facilities in government and university buildings, which were overwhelmingly clustered within a single audit area and may differ contextually from other facilities. We re-calculated MFPT Index and sub-domain scores (1) *excluding* government and university buildings and (2) *restricted to* government and university buildings. We then compared scores using t-tests and re-examined differences between funding types within the reduced sample (excluding government and university buildings). Sensitivity analyses did not reveal significant differences (see Supplementary Material). All analyses were conducted using R (version 4.3.1).

## Results

We audited 125 women’s, gender neutral, and family bathroom facilities across 112 unique locations in Atlanta, Georgia. Of these locations, 57.1% (n=64) were privately funded and 42.9% (n=48) were government-funded (Table 1). Location type varied, with chain restaurants or coffee shops representing the largest category (21.4%, n=24), followed by universities (19.6%, n=22), park or recreation centers (11.6%, n=13), and big-box retail stores (8.9%, n=10). Additional locations included government buildings, libraries, grocery stores or markets, tourist attractions, health facilities, transit stations, pharmacies, shopping malls or multipurpose buildings, gas stations, and places of worship.

**Table 1.** Descriptive statistics of audited locations (n=112)

| **Characteristic** | **n (%)** |
|---|---|
| **Funding** |  |
| Privately funded | 64 (57.1) |
| Government funded | 48 (42.9) |
| **Location type** |  |
| Chain restaurant or coffee shop | 24 (21.4) |
| University | 22 (19.6) |
| Park or recreation center | 13 (11.6) |
| Big-box retail store | 10 (8.9) |
| Government building | 5 (4.5) |
| Library | 4 (3.6) |
| Grocery store, convenience store, or market | 8 (7.1) |
| Tourist attraction | 7 (6.3) |
| Health facility | 6 (5.4) |
| Transit station | 3 (2.7) |
| Pharmacy | 3 (2.7) |
| Shopping mall or multipurpose building | 3 (2.7) |
| Gas station | 2 (1.8) |
| Place of worship (e.g., church) | 2 (1.8) |
| **Toilet location** |  |
| In a building | 105 (93.8) |
| Standalone bathroom facility | 7 (6.3) |
| **Open 24/7** |  |
| No | 88 (78.6) |
| Yes | 6 (5.4) |
| Unclear | 18 (16.1) |
| **Need permission to use** | 22 (19.6) |
| **Need to access building via turnstile or gate** | 16 (14.3) |
| **At least one wheelchair accessible entrance** | 103 (92.0) |

The vast majority of bathrooms were located within a building (93.8%, n=105), with relatively few standalone bathroom facilities identified (6.3%, n=7). Only six locations appeared to operate 24 hours a day, seven days a week, while hours of operation were unclear for 16.1% (n=18) of locations. Some locations required permission to access bathrooms (19.6%, n=22) or entry through a turnstile or gate (14.3%, n=16). Most locations (92.0%, n=103) had at least one wheelchair-accessible entrance.

Among the 125 bathroom facilities, 65.6% (n=82) were designated for women, 21.6% (n=27) were gender-neutral, and 12.8% (n=16) were family bathrooms or had no gender designation (Table 2). Bathroom facility capacity varied: 53.6% (n=67) were multi-occupant bathrooms with stalls, 43.2% (n=54) of facilities were single-occupant bathrooms without stalls, and 3.2% (n=4) were single-occupant bathrooms with stalls. On average, facilities had 2.7 toilets (standard deviation [SD]=2.3). However, due to prolonged occupancy of some stalls (>10 minutes), an average of 2.5 toilets per facility were assessed, yielding data on 310 unoccupied toilets across all facilities.

**Table 2.** Select characteristics of audited bathroom facilities (n=125)

| **Characteristics** | **n (%)** |
|---|---|
| **Facility gender designation** |  |
| Women’s | 82 (65.6) |
| Gender neutral | 27 (21.6) |
| No gender specified/family | 16 (12.8) |
| **Facility capacity** |  |
| Multi-occupant, stalls | 67 (53.6) |
| Single-occupant, no stalls | 54 (43.2) |
| Single-occupant, stalls | 4 (3.2) |
| **Number of toilets (mean (SD))** | 2.7 (2.3) |
| **Number of unoccupied toilets (mean (SD))** | 2.5 (2.2) |

SD: Standard deviation

### Overall and sub-domains of MFPT

Table 3 presents MFPT Index and sub-domain scores overall and by funding type. The mean MFPT Index score across all facilities (n=125) was 0.797, indicating that facilities met approximately 80% of MFPT criteria. Sub-domain scores varied considerably, ranging from 0.974 for cleanliness to 0.490 for availability of MHH resources.

**Table 3.** Descriptive statistics of Menstrual Friendly Public Toilet sub-domains by funding type (n=125)

|  | Total | Government-funded | Privately funded |
|---|---|---|---|
| **Number of bathroom facilities** | 125 | 54 | 71 |
| **Overall MFPT Index Scores** | **0.797** | **0.808** | **0.789** |
| **Sub-domain 1: Accessibility scores** | **0.622** | **0.611** | **0.631** |
|  | n (%) | n (%) | n (%) |
| No permission needed to use the toilet | 100 (80.0) | 44 (81.5) | 56 (78.9) |
| Visible sign indicating toilet entrance | 57 (45.6) | 28 (51.9) | 29 (40.8) |
| No prior access to an establishment needed | 106 (84.8) | 40 (74.1)* | 66 (93.0)* |
| Open 24/7 | 7 (5.6) | 2 (3.7) | 5 (7.0) |
| No fee | 119 (95.2) | 51 (94.4) | 68 (95.8) |
| **Sub-domain 2: Structural privacy/safety scores** | **0.954** | **0.947** | **0.964** |
|  | n (%) | n (%) | n (%) |
| Functional entrance door or no door by design | 125 (100) | 54 (100) | 71 (100) |
| ≥75% unoccupied stalls with functional doors | 69 (97.2)<sup>^</sup> | 39 (97.5)<sup>+</sup> | 30 (96.8)<sup>#</sup> |
| ≥75% of unoccupied stalls with functional locks | 64 (90.1)<sup>^</sup> | 34 (85.0)<sup>+</sup> | 30 (96.8)<sup>#</sup> |
| Functional electric light | 125 (100) | 54 (100) | 71 (100) |
| **Sub-domain 3: Structure scores** | **0.930** | **0.952\*** | **0.913\*** |
|  | n (%) | n (%) | n (%) |
| ≥75% of unoccupied facilities or stalls with functional hooks/shelves | 87 (69.6) | 44 (81.5)* | 43 (60.6)* |
| Presence of windows, skylights, or electric light | 125 (100) | 54 (100) | 71 (100) |
| Good or adequate condition toilets | 124 (99.2) | 54 (100) | 70 (98.6) |
| Functional sink(s) | 123 (98.4) | 53 (98.1) | 70 (98.6) |
| Ventilation | 122 (97.6) | 52 (96.3) | 70 (98.6) |
| **Sub-domain 4: Availability of resources scores** | **0.899** | **0.878** | **0.915** |
|  | n (%) | n (%) | n (%) |
| ≥75% of unoccupied facilities or stalls with at least 1 roll of toilet paper | 123 (98.4) | 53 (98.1) | 70 (98.6) |
| Available and useable trash cans/bins | 117 (93.6) | 49 (90.7) | 68 (95.8) |
| ≥75% of unoccupied facilities or stalls with functional toilet seats | 122 (97.6) | 51 (94.4) | 71 (100) |
| Water available in facility | 124 (99.2) | 53 (98.1) | 71 (100) |
| Ratio ≥0.5 functional sinks per number of toilets | 101 (80.8) | 43 (79.6) | 58 (81.7) |
| Ratio ≥0.5 soap/liquid dispensers/hand sanitizer per number of toilets | 82 (65.6) | 32 (59.3) | 50 (70.4) |
| At least one functional hand-dryer or stocked towels | 118 (94.4) | 51 (94.4) | 67 (94.4) |
| **Sub-domain 5: Cleanliness scores** | **0.974** | **0.972** | **0.975** |
|  | n (%) | n (%) | n (%) |
| Visibly clean | 112 (89.6) | 48 (88.9) | 64 (90.1) |
| No or some noticeable odor | 125 (100) | 54 (100) | 71 (100) |
| Good or adequately clean floors | 125 (100) | 54 (100) | 71 (100) |
| Good or adequate privacy provided by walls/ roof coverage | 125 (100) | 54 (100) | 71 (100) |
| **Sub-domain 6: Availability of MHH resources scores** | **0.490** | **0.537** | **0.454** |
|  | n (%) | n (%) | n (%) |
| Mirror adequate for checking clothes | 118 (94.4) | 50 (92.6) | 68 (95.8) |
| ≥75% of unoccupied facilities and stalls with disposal bins | 72 (57.6) | 37 (68.5)* | 35 (49.3)* |
| ≥75% of unoccupied facilities and stalls with lined disposal bins | 50 (40.0) | 24 (44.4) | 26 (36.6) |
| ≥75% of unoccupied facilities and stalls with lidded disposal bins | 61 (48.8) | 31 (57.4) | 30 (42.3) |
| Products stocked or working machine | 5 (4.0) | 3 (5.6) | 2 (2.8) |

\*Statistically significant at p < 0.05, ^n=71, +n=40, #n=31

Bathroom facilities performed consistently well across four sub-domains – structural privacy/safety, structure, availability of resources, and cleanliness – with average scores approaching or exceeding 0.90. Within the structural privacy/safety sub-domain, all facilities had either a functional entrance doors or were designed without one and had functional electric lighting. Among facilities with stalls, 97.2% (n=69/71) had functional stall doors and 90.1% (n=64/71) had functional stall locks. Within the structure sub-domain, all or nearly all facilities had adequate lighting (100%, n=125), toilets in good or adequate condition (99.2%, n=124), functional sinks (94.8%, n=123), and ventilation (97.6%, n=122). However, fewer facilities (69.6%, n=87) had functional hooks or shelves in stalls. For general resource availability (sub-domain 4), most facilities had toilet paper (98.4%, n=123), trash bins (93.6%, n=117), functional toilet seats (97.6%, n=122), access to water (99.2%, n=124), and hand-drying options (94.4%, n=118). However, some facilities did not meet recommended ratios of sinks (19.2%, n=24) or soap/hand hygiene dispensers (34.4%, 43) relative to the number of toilets. Lastly, cleanliness scores were uniformly high. All facilities met the criteria for minimal odor, clean or adequately clean floors, and sufficient privacy from walls and roof. Most facilities (89.6%, n=112) were also visibly clean.

In contrast, accessibility (sub-domain 1) and availability of MHH resources (sub-domain 6) had the lowest scores (0.622 and 0.490, respectively). Fewer than half of facilities (45.6%, n=57) had visible signage indicating bathroom entrances and only 5.6% (n=7) were open 24 hours per day. However, most facilities did not require payment (95.2%, n=119), permission (80.0%, n=100), or prior patronage/access (84.8%, n=106). Availability of MHH resources was particularly limited. Only 4.0% (n=5) of facilities had menstrual materials or functional dispensers. Disposal bins were also frequently inadequate: 42.4% (n=53) lacked MHH-specific disposal bins, 60.0% (n=75) lacked lined disposal bins, and 51.2% (n=64) lacked lidded disposal bins in stalls. However, 94.4% (n=118) of facilities had mirrors adequate for checking clothes.

### Overall and sub-domains of MFPT by funding

Overall MFPT Index and sub-domain scores did not differ significantly by funding type. The only statistically significant difference was observed in the structure sub-domain: government-funded facilities had higher scores (0.952) than privately funded facilities (0.913; p=0.027), although both were relatively high. At the item level, three indicators differed significantly. Privately funded facilities had unrestricted building access more often than government-funded facilities (93.0% vs. 74.1%, p=0.008). In contrast, government-funded facilities had functional hooks/shelves (81.5% vs. 60.6%, p=0.020) and MHH-specific disposal bins (68.5% vs. 49.3%, p=0.049) more often than privately funded facilities.

### Overall and sub-domains of MFPT by neighborhood

Among the sub-sample of bathroom facilities in business, residential, tourist, and transit hub station neighborhoods (n=57), the overall MFPT Index score was slightly higher than in the total sample (0.817 vs. 0.797, Table 4). Bathroom facilities in the tourist area scored the highest (0.862), followed by those in the business area (0.838), transit hub station (0.811), and residential area (0.748). Sub-domain scores overall and across neighborhoods followed similar trends to those seen in the overall sample: facilities performed consistently well across structural privacy/safety, structure, availability of resources, and cleanliness sub-domains with average scores approaching or exceeding 0.90. In contrast, accessibility (sub-domain 1) and availability of MHH resources (sub-domain 6) had the lowest scores overall and across neighborhoods.

**Table 4.** Descriptive statistics of Menstrual Friendly Public Toilet sub-domains by neighborhood type (n=57)

|  | Total | Business area | Residential area | Tourist area | Transit hub station |
|---|---|---|---|---|---|
| **Number of bathroom facilities** | 57 | 32 | 5 | 8 | 12 |
| **Overall MFPT Index Scores** | **0.817** | **0.838** | **0.748** | **0.862** | **0.811** |
| **Sub-domain 1: Accessibility scores** | **0.639** | **0.638** | **0.680** | **0.700** | **0.583** |
|  | n (%) | n (%) | n (%) | n (%) | n (%) |
| No permission needed to use the toilet | 46 (80.7) | 27 (84.4) | 4 (80.0) | 8 (100.0) | 7 (58.3) |
| Visible sign indicating toilet entrance | 33 (57.9) | 20 (62.5) | 3 (60.0) | 4 (50.0) | 6 (50.0) |
| No prior access to an establishment needed | 47 (82.5) | 23 (71.9) | 5 (100.0) | 8 (100.0) | 11 (91.7) |
| Open 24/7 | 1 (1.8) | 1 (3.1) | 0 (0.0) | 0 (0.0) | 0 (0.0) |
| No fee | 55 (96.5) | 31 (96.9) | 5 (100.0) | 8 (100.0) | 11 (91.7) |
| **Sub-domain 2: Structural privacy/safety scores** | **0.951** | **0.955** | **0.875** | **0.953** | **0.950** |
|  | n (%) | n (%) | n (%) | n (%) | n (%) |
| Functional entrance door or no door by design | 57 (100.0) | 32 (100.0) | 5 (100.0) | 8 (100.0) | 12 (100.0) |
| ≥75% unoccupied stalls with functional doors | 35 (97.2)<sup>^</sup> | 21 (95.5)<sup>+</sup> | 1 (100.0)<sup>#</sup> | 8 (100.0) | 5 (100.0)<sup>@</sup> |
| ≥75% of unoccupied stalls with functional locks | 33 (97.1)<sup>~</sup> | 20 (90.9)<sup>+</sup> | 1 (100.0)<sup>#</sup> | 8 (100.0) | 4 (80.0)<sup>@</sup> |
| Functional electric light | 57 (100.0) | 32 (100.0) | 5 (100.0) | 8 (100.0) | 12 (100.0) |
| **Sub-domain 3: Structure scores** | **0.919** | **0.950** | **0.840** | **0.900** | **0.883** |
|  | n (%) | n (%) | n (%) | n (%) | n (%) |
| ≥75% of unoccupied facilities or stalls with functional hooks/shelves | 38 (66.7) | 26 (81.2) | 1 (20.0) | 4 (50.0) | 7 (58.3) |
| Presence of windows, skylights, or electric light | 57 (100.0) | 32 (100.0) | 5 (100.0) | 8 (100.0) | 12 (100.0) |
| Good or adequate condition toilets | 56 (98.2) | 32.0 (100.0) | 5 (100.0) | 8 (100.0) | 11 (91.7) |
| Functional sink(s) | 56 (98.2) | 32 (100.0) | 5 (100.0) | 8 (100.0) | 11 (91.7) |
| Ventilation | 55 (96.5) | 30 (93.8) | 5 (100.0) | 8 (100.0) | 12 (100.0) |
| **Sub-domain 4: Availability of resources scores** | **0.912** | **0.906** | **0.943** | **0.875** | **0.940** |
|  | n (%) | n (%) | n (%) | n (%) | n (%) |
| ≥75% of unoccupied facilities or stalls with at least 1 roll of toilet paper | 56 (98.2) | 31 (96.9) | 5 (100.0) | 8 (100.0) | 12 (100.0) |
| Available and useable trash cans/bins | 53 (93.0) | 28 (87.5) | 5 (100.0) | 8 (100.0) | 12 (100.0) |
| ≥75% of unoccupied facilities or stalls with functional toilet seats | 57 (100.0) | 32 (100.0) | 5 (100.0) | 8 (100.0) | 12 (100.0) |
| Water available in facility | 57 (100.0) | 32 (100.0) | 5 (100.0) | 8 (100.0) | 12 (100.0) |
| Ratio ≥0.5 functional sinks per number of toilets | 46 (80.7) | 28 (87.5) | 4 (80.0) | 4 (50.0) | 10 (83.3) |
| Ratio ≥0.5 soap/liquid dispensers/hand sanitizer per number of toilets | 40 (70.2) | 21 (65.6) | 5 (100.0) | 5 (62.5) | 9 (75.0) |
| At least one functional hand-dryer or stocked towels | 55 (96.5) | 31 (96.9) | 4 (80.0) | 8 (100.0) | 12 (100.0) |
| **Sub-domain 5: Cleanliness scores** | **0.978** | **0.984** | **0.950** | **0.969** | **0.979** |
|  | n (%) | n (%) | n (%) | n (%) | n (%) |
| Visibly clean | 52 (91.2) | 30 (93.8) | 4 (80.0) | 7 (87.5) | 11 (91.7) |
| No or some noticeable odor | 57 (100.0) | 32 (100.0) | 5 (100.0) | 8 (100.0) | 12 (100.0) |
| Good or adequately clean floors | 57 (100.0) | 32 (100.0) | 5 (100.0) | 8 (100.0) | 12 (100.0) |
| Good or adequate privacy provided by walls/ roof coverage | 57 (100.0) | 32 (100.0) | 5 (100.0) | 8 (100.0) | 12 (100.0) |
| **Sub-domain 6: Availability of MHH resources scores** | **0.572** | **0.594** | **0.200** | **0.775** | **0.533** |
|  | n (%) | n (%) | n (%) | n (%) | n (%) |
| Mirror adequate for checking clothes | 56 (98.2) | 31 (96.9) | 5 (100.0) | 8 (100.0) | 12 (100.0) |
| ≥75% of unoccupied facilities and stalls with disposal bins | 38 (66.7) | 23 (71.9) | 0 (0.0) | 8 (100.0) | 7 (58.3) |
| ≥75% of unoccupied facilities and stalls with lined disposal bins | 33 (57.9) | 21 (65.6) | 0 (0.0) | 6 (75.0) | 6 (50.0) |
| ≥75% of unoccupied facilities and stalls with lidded disposal bins | 34 (59.6) | 20 (62.5) | 0 (0.0) | 7 (87.5) | 7 (58.3) |
| Products stocked or working machine | 2 (3.5) | 0 (0.0) | 0 (0.0) | 2 (25.0) | 0 (0.0) |

^n=36, ~n=34, +n=22, #n=1, @n=5

## Discussion

Having safe, conveniently located, private, and properly resourced bathrooms in public spaces is critical for MHH, participation in public space overall, and participation in public activities (3–7). Publicly accessible bathroom facilities in Atlanta, Georgia demonstrated high menstrual friendliness, with MFPT scores comparable to or exceeding those observed in other global cities. Facilities averaged an MFPT Index score of 0.797, indicating that they met approximately 80% of criteria, which is comparable to facilities in Barcelona (0.749), New York City (0.725), and Osaka (0.793) (14). These findings suggest that a high-income US city can meet baseline menstrual-friendly criteria. However, the relatively high scores mask important gaps in accessibility and MHH resources, indicating that adequacy in infrastructure does not necessarily translate to equitable access or meeting the needs of those who menstruate. Furthermore, these MFPT findings should be interpreted alongside broader patterns of public bathroom availability in Atlanta, which revealed that (1) over half of identified locations were not accessible to the public, even when applying an inclusive definition of “public” facilities; and (2) the average density of publicly accessible bathrooms for any gender varied across areas (range: 2.55-76.4 per km^2^), highlighting spatial inequities in access (29). By applying the MFPT framework in Atlanta, this study extends the global evidence base and demonstrates that MHH inequities persist across diverse economic and urban settings.

Despite strong performance on privacy/safety, structure, general resources, and cleanliness indicators, key gaps remained in accessibility and MHH resource provision, mirroring patterns observed in urban settings globally (14). Notably, only seven audited bathrooms were open 24 hours a day, seven days a week and over half did not have visible signs indicating a bathroom entrance. These findings highlight not only limited availability but also barriers to navigability and predictability in accessing sanitation. This is further reinforced by broader patterns of access across Atlanta, where more than half of identified locations were not publicly accessible despite being located in high-traffic areas such as tourist destinations and university campuses (29). For individuals managing menstruation, uncertainty about whether a bathroom is open and where it is located can significantly constrain mobility and participation in public space (17, 18, 21).

These barriers are particularly salient for populations with fewer alternatives, including people experiencing houselessness, people with health conditions, and workers without flexible schedules (7, 9, 17, 21). While public bathrooms may not be economically feasible to be established and maintained everywhere, strategic investments should prioritize both spatial coverage and consistent accessibility (e.g., extended hours, clear signage, reliable access conditions).

The availability of MHH resources emerged as the most consistent and substantial gap, highlighting how little attention is paid to menstrual management needs in public spaces. Aligned with existing research (14), this sub-domain scored the lowest across facilities and the availability of menstrual materials or products was very uncommon. Some US cities have begun requiring free menstrual materials or products in public bathrooms (e.g., Ann Arbor, Michigan (33); Cleveland, Ohio (34)), which can help improve the menstrual friendliness of facilities. However, disposal bins remain a significant, unaddressed issue. Together, these gaps point to a broader pattern in which MHH needs are treated as discretionary rather than essential components of public sanitation infrastructure. Without addressing cultural stigma and taboos around menstruation, as well as the absence of policies focused on MHH in the public domain, such resources will continue to be de-prioritized in planning and funding.

Differences in MFPT Index scores across neighborhood types demonstrate how urban planning can shape access to menstrual-friendly facilities, with residential areas being consistently underserved in Atlanta and urban settings across the world. Similar to the multi-city MFPT study (14), we found that facilities in the tourist area had the highest MFPT scores while those in the residential area had the lowest. This represents greater investment, visibility, and maintenance in income generating neighborhoods where visitors frequent and an underinvestment in public goods in residential areas, suggesting that residents may face less accessible and lower quality facilities compared to visitors. This pattern reflects broader urban investment dynamics in which resources are concentrated in high-visibility, revenue-generating areas while everyday residential environments receive less attention (35–38). Public facilities in residential neighborhoods need to be equally prioritized to ensure that sanitation access supports daily life, not just tourism or economic activity.

The lack of significant differences by funding type suggests that both government-funded and privately funded bathrooms contribute to uneven sanitation quality, though with distinct structural trade-offs as several facility characteristics varied. Privately funded facilities were less likely to include features such as hooks or shelves, adequate lighting, well-maintained toilets, functional seats, and ventilation. This aligns with findings from our broader study examining non-MHH characteristics (29), suggesting that while private-sector provision can expand access, it does not consistently ensure quality or user-centered design. These findings highlight a fragmented “hybrid” sanitation system in which responsibility is distributed across public and private actors without consistent standards or accountability (8, 9, 12). As traditional public bathrooms continue to decline and reliance on private spaces increases, these trade-offs have important implications for equitable access and user experience.

### Limitations

Findings from this study should be interpreted in light of several limitations. First, the study was cross-sectional and reflects conditions at a single point in time. Specifically, facilities were only audited during weekday, non-holiday business hours and availability may differ during evenings, weekends, or holidays. Longitudinal audits could strengthen understanding of temporal variability in sanitation access. Second, although we audited 15 areas across Atlanta, facilities in non-sampled areas may differ and site selection was informed by the research team’s local knowledge, which may limit generalizability. Relying on local knowledge rather than a randomized spatial sampling strategy could inadvertently introduce selection bias based on the selection of familiar or high traffic areas. As a result, our findings may inadvertently overestimate the true baseline of menstrual-friendly sanitation availability. However, leveraging local knowledge to include areas where public sanitation demand is the highest, such as high-pedestrian corridors, transit hubs, and areas frequented by vulnerable populations, was a distinct advantage in this study. Lastly, this study focused on publicly accessible facilities and did not capture user experiences or perceptions of bathroom quality, which are also critical components of menstrual management in public spaces.

## Conclusion

While Atlanta’s public bathrooms meet many baseline standards of menstrual friendliness, persistent gaps in accessibility and MHH resource provision highlight that adequacy in infrastructure does not necessarily translate to equitable access. These findings point to critical needs across infrastructure, resource provision, programming, and policy. From a facility design and management perspective, relatively low-cost improvements – such as visible entrance signs, sufficient soap dispensers, hooks or shelves, appropriate in-stall disposal bins, and the provision of menstrual materials – could substantially improve menstrual friendliness. However, addressing infrastructure and resource provision alone is insufficient. Efforts to reduce cultural stigma surrounding menstruation, particularly in public spaces, are also needed to ensure that MHH needs are prioritized in planning and resource allocation. Lastly, at the systems level, strategic policy and investment are required. While policies mandating free menstrual materials in public facilities represent an important step, broader and more comprehensive approaches are needed. These include establishing standardized criteria for menstrual-friendly public facilities, allocating resources to residential neighborhoods, and ensuring consistent accessibility and quality across both government and privately managed spaces. As reliance on privately operated facilities grows and government-funded bathrooms decline, coordinated, equity-focused approaches to urban sanitation will be essential for supporting participation in public life and advancing MHH equity.

## Data Availability

All relevant data for this study are publicly available from the OSF repository (https://osf.io/fm9by).

## Acknowledgements

We are grateful to the Menstrual Friendly Public Toilet (MFPT) Working Group, particularly Angela Nguyen, Marni Sommer, and Anderew Maroko, for sharing their resources and experiences conducting public toilet audits.

## Supporting information captions

**S1 Table.** Sensitivity analysis of MFPT Index and Sub-domain scores

**S2 Table.** Sensitivity analysis of MFPT index by funding source

